# Measuring the policy citation rate of United Kingdom (UK) paramedic and ambulance service research: a bibliometric evaluation using BMJ Impact Analytics and the *amber* repository

**DOI:** 10.64898/2026.09.05.26362342

**Authors:** Matt Holland

## Abstract

**Context/Objective:** Demonstrating real-world impact is increasingly important for prehospital researchers. While biomedical sciences show high altmetric coverage, policy citation density in paramedicine remains largely unknown. This study evaluates the policy citation rate of UK ambulance service research, measures uptake differences between clinical practice guidelines and broader health policies and maps its geographic dissemination.

**Methods:** Following the BiBLio reporting guidelines, all publications with a Digital Object Identifier (DOI) where at least one author was working in an NHS ambulance service were extracted from the *amber* repository covering 2006 to 2026 (N=2,095). Policy and clinical guideline citations were tracked using BMJ Impact Analytics. Document classifications and issuing organisation metadata were analysed in Microsoft Excel using descriptive bibliometrics.

**Key Findings:** The overall policy citation rate was 23.48% (n=492/2,095), with 53.66% of cited papers cited once and 1.63% achieving 10 or more citations. A total of 1,143 citations were identified across 760 clinical practice guidelines (66.49%) and 383 health policies (33.51%), producing a guideline-to-policy ratio of 1.98:1. Multicentre clinical trials accrued high scholarly and policy citation volumes, whereas service design and workforce studies achieved high policy translation despite modest scholarly citations. Of 704 citations with resolvable geographic metadata, 46.45% (n=327) were domestic UK citations, 32.67% (n=230) originated from international Anglophone jurisdictions, 13.07% (n=92) from non-Anglophone European nations, and 4.83% (n=34) from international bodies.

**Conclusion/Significance:** UK paramedic and ambulance service research demonstrates measurable translation into clinical practice guidelines and health policy, although its 23.48% policy citation rate places it towards the lower end of comparable empirical healthcare portfolios. The disproportionate uptake in clinical guidelines highlights the reliance of guideline panels on high-quality prehospital evidence. Maximising translational reach requires active dissemination by embedded researchers, explicit practice recommendations within publications, and systematic green open access preservation via sector repositories such as *amber*.

## Background

Researchers are increasingly being asked to demonstrate the real-world impact of their research. A possible measure is to look at the citation of research outputs in policy documents and guidelines. Within the area of medicine and related clinical topics, the inclusion of research in policies and clinical guidelines broadly indicates the translation of evidence into clinical practice and patient care.

Early studies have looked at policies included as part of the altmetric suite (Fang et al., 2020). Fang et al. note in this study that Biomedical and Health Sciences were among the topics that had the highest altmetric coverage. In another study, Tattersall & Carroll (2018) urged caution when relying on altmetric data alone, identifying challenges in accurately mining and identifying unstructured citations in policy documents.

The development of new, dedicated policy databases such as Overton and BMJ Impact Analytics now enables policy citation analysis with greater accuracy and a substantial body of data. Using this infrastructure, research by Szomszor and Adie (2022) confirmed that health sciences and medicine account for the largest overall volume of policy document citations, establishing a robust source for evaluating translational impact.

Work in this area frequently approaches analysis using discrete datasets to test translational impact within specific subject domains (Bornmann et al., 2022; Llewellyn et al., 2026; Zachariah & Bond, 2026), the outputs of defined research organisations or groups (Tattersall & Carroll, 2018; Tunn et al., 2025; Omondi et al., 2026; van Elsland, 2024), or defined geographical areas and repository collections (McManus et al., 2025).

Unlike other areas of bibliometric research, there are no established subject benchmarks against which to measure citation performance in policy and guidelines. Szomszor & Adie (2022) demonstrated that medicine contains sufficient citation density to support subject-level benchmarking, comparative analysis still relies on baselines derived from individual studies against which to measure translational impact.

### Current Evidence Gap

Studies of research impact that encompass a multidisciplinary corpus of literature typically generate a low translational effect. Table 1 summarises a range of studies that illustrate this. Studies that used the policy strand of Altmetric.com—acknowledged to be sparse—report the lowest measures (0.32%--1.41%). Studies using Overton, with its dedicated, full-text policy database, report comparatively higher citation rates (3.90%--5.86%).

**Table 1.** Broad Multidisciplinary & General Scientific Baselines.

| Study | Database | Corpus & Scope | Policy Citation Rate (% / Count) |
| --- | --- | --- | --- |
| Haunschild & Bornmann (2017) | Altmetric.com | 11.3M Web of Science publications (all disciplines) | 0.32% of publications |
| Tattersall & Carroll (2018) | Altmetric.com | 96,550 University of Sheffield outputs (institutional, all disciplines) | 1.41% (1,463 / 96,550 outputs) |
| Szomszor & Adie (2022) | Overton | 4.85M global policy PDFs (cross-disciplinary policy corpus) | 11.76% of policy PDFs cited academic literature (570,830 PDFs) |
| Bornmann et al. (2022) | Overton & Scopus | Scopus-indexed research items with a DOI (global scientific baseline) | 4.98% of Scopus DOI items |
| Fang et al. (2024) | Overton & Web of Science | 18M Web of Science publications, 2010–2019 (all disciplines) | 3.90% of publications cited in ≥ 1 policy document |
| McManus et al. (2025) | Overton | 615,633 publications from São Paulo, Brazil (regional multidisciplinary corpus) | 5.86% (36,100 / 615,633 documents) |

Szomszor & Adie (2022) demonstrated that biological sciences and medicine among other disciplines have a high translational effect, in that published research in these areas is more likely to cited in guidelines and policies. We would expect then that corpuses of research literature in specific subject areas like healthcare research will have a higher translational effect. This is broadly supported by the evidence form recent studies set out in Table 2.

**Table 2.** Targeted Health, Biomedical & Applied Research Portfolios.

| Study | Database | Corpus & Scope | Policy Citation Rate (% / Count) |
| --- | --- | --- | --- |
| Llewellyn et al. (2023) | Overton | 134,607 NIH Clinical & Translational Science Awards (CTSA) papers | 14.6% (19,673 / 134,607); 25% for substance abuse research |
| Omondi et al. (2026) | Overton | 10,267 articles using Demographic and Health Surveys (DHS) data | 29.9% (3,073 / 10,267 articles) |
| Tunn et al. (2025) | Overton | 110 articles from Policy Research Unit in Maternal and Neonatal Health | 37.3% (41 / 110 articles; reported as ~40%) |
| van Elsland et al. (2024) | Overton | Imperial College COVID-19 Response Team (ICCRT) outputs | 43.0% of research outputs cited across 41 countries |
| Zachariah & Bond (2026) | Overton | 728 articles in International Journal of Pharmacy Practice (20-year archive) | 54.1% (394 / 728 articles) |

While these provide an empirical range from 14.6% to 54.1% for a Policy citation rate there are clearly factors within each study that might influence the transitional effect. We might speculate that this would include a strong subject focus on a clinical topic.

This study looks specifically at paramedic and ambulance service research produced in the United Kingdom. The primary aim is to determine the translational effect and policy citation rate establishing how it benchmarks against existing multidisciplinary and healthcare-specific literature. Specifically:

1. Determine the overall policy citation rate and translational volume across the UK ambulance service research represented by the contents of the amber repository;
2. Measure the difference in uptake between clinical practice guidelines and broad health policy citations; and
3. Characterise the geographic spread and international reach of health policy and clinical guidelines referencing UK paramedic and ambulance service research.

## Methods

This study is reported in accordance with the preliminary guideline for reporting bibliometric reviews of the biomedical literature (BIBLIO) checklist (Montazeri et al., 2023). The completed BIBLIO checklist is provided in Supplementary File 1. Data were extracted from amber [https://amber.openrepository.com] in June 2026. Amber is the United Kingdom (UK) repository for NHS ambulance services (11 ambulance services in England, Scottish Ambulance Service, Welsh Ambulance Service and Northern Ireland Ambulance Service). amber contains research where one author is a member of an NHS ambulance service covering the period from 2006 to 2026.

From this data we further extracted all documents with a Digital Object Identifier (DOI). The Isle of Wight NHS Trust had no outputs with a DOI in the repository at the time of extraction. Duplicate DOIs were removed. The final data set contained N=2098 outputs. Three erroneous DOI’s were removed.

Using the DOIs we ran searches for each ambulance service in BMJ Impact Analytics - a database of policy documents for healthcare and clinical guidelines to aid decisions on patient care and diagnosis. The results were exported and augmented with additional document-level data: a separate citation count for Guidelines and Policies and the geographic origin of the policy or guideline. All data were compiled in Microsoft Excel to perform descriptive bibliometric analysis.

## Results

The policy citation rate is summarised in Table 3. The overall rate for UK paramedic and ambulance service research is 23.48% (n=492/2,095) where BMJ Impact Analytics has identified at least one reference in a healthcare policy or clinical guideline. Over half are cited just once 53.66% (n=264) with over a quarter achieving a relatively high citation rate of three or more citations. There is a small group of 8 papers, 1.63% of the DOI’s searched for and 0.38% of the whole corpus that achieved 10 or more citations.

**Table 3.**
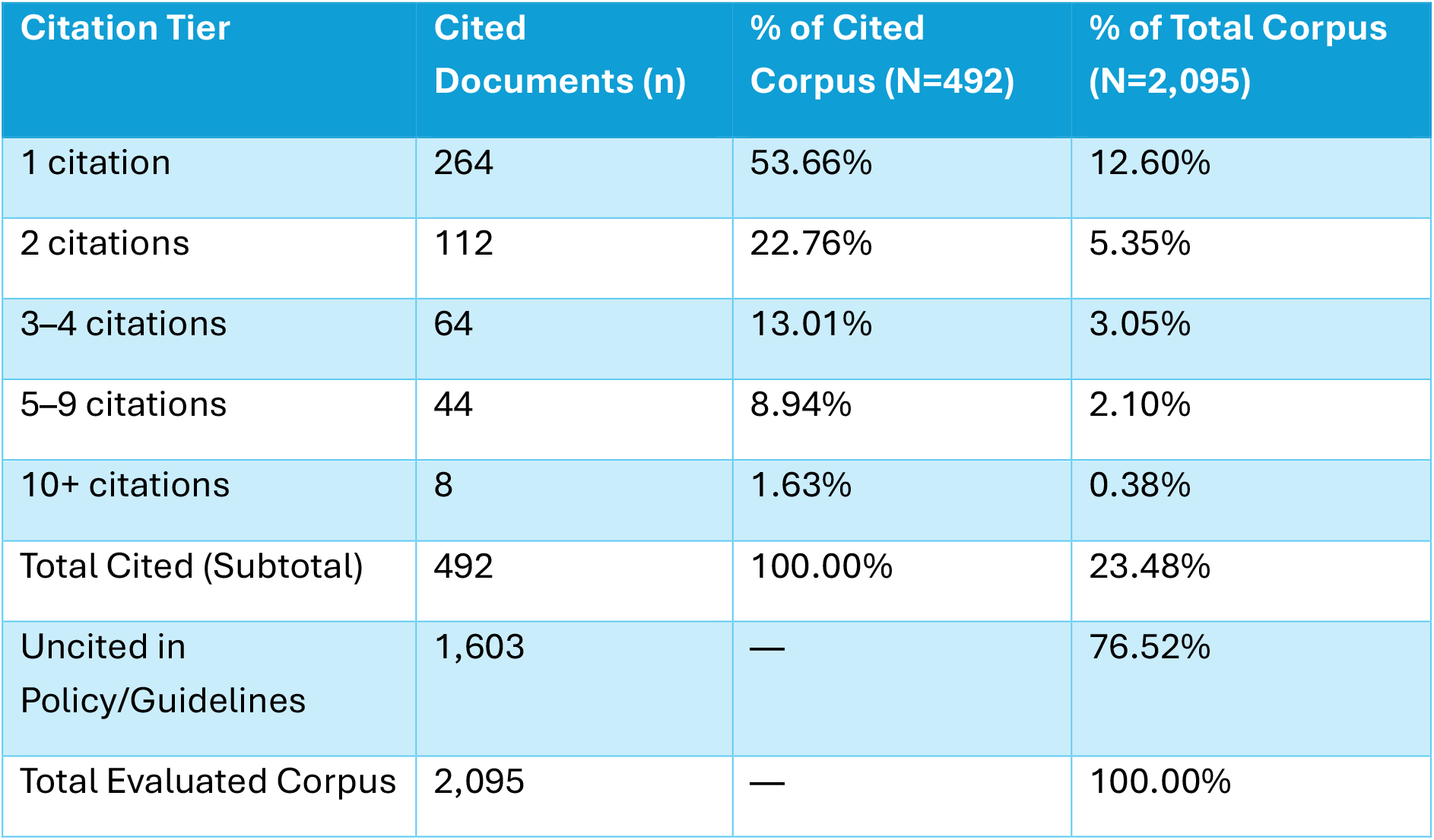
Policy Citation Rates for paramedic and ambulance research in the UK 2006 – 2026.

| Citation Tier | Cited Documents (n) | % of Cited Corpus (N=492) | % of Total Corpus (N=2,095) |
| --- | --- | --- | --- |
| 1 citation | 264 | 53.66% | 12.60% |
| 2 citations | 112 | 22.76% | 5.35% |
| 3–4 citations | 64 | 13.01% | 3.05% |
| 5–9 citations | 44 | 8.94% | 2.10% |
| 10+ citations | 8 | 1.63% | 0.38% |
| Total Cited (Subtotal) | 492 | 100.00% | 23.48% |
| Uncited in Policy/Guidelines | 1,603 | — | 76.52% |
| Total Evaluated Corpus | 2,095 | — | 100.00% |

Citation patterns across the top eight publications (Table 4) illustrate distinct translational patterns. All eight papers generated high overall academic and policy citations, randomised controlled trials and major multicentre clinical investigations (e.g., AIRWAYS-2 and PARAMEDIC-2) achieved both high citation scores and high policy uptake. Service design, workforce studies and triage pathways achieve a high translational effect reflected in a high policy citation rate with lower citation scores.

**Table 4.** Top 8 Highly Cited Publications (10+ Citations)

| # | Title | Journal | BMJ IA | Dimensions / Plum X |
| --- | --- | --- | --- | --- |
| 1 | Impact on Clinical and Cost Outcomes of a Centralized Approach to Acute Stroke Care | PLOS ONE | 25 | 102 |
| 2 | Epidemiology and outcomes from out-of-hospital cardiac arrests in England | Resuscitation | 15 | 316 |
| 3 | Effect of a Strategy of a Supraglottic Airway Device vs Tracheal Intubation During Out-of-Hospital Cardiac Arrest | JAMA | 15 | 429 |
| 4 | Effectiveness of paramedic practitioners in attending 999 calls from elderly patients | BMJ | 14 | 311 |
| 5 | Feasibility of an Ambulance-Based Stroke Trial, and Safety of Glyceryl Trinitrate | Stroke | 12 | 146 |
| 6 | Does Use of the Recognition Of Stroke In the Emergency Room Stroke Assessment Tool Enhance Stroke Recognition by Ambulance Clinicians? | Stroke | 12 | 67 |
| 7 | A Randomized Trial of Epinephrine in Out-of-Hospital Cardiac Arrest | New England Journal of Medicine | 11 | 722 |
| 8 | Temporal and geographic patterns of stab injuries in young people: a retrospective cohort study | BMJ Open | 10 | 42 |

Citations originating from clinical practice guidelines substantially exceeded those from broader health policy documents, as shown in Table 5. Across all identified citations (n=1,143), nearly twice as many appeared in clinical guidelines (66.49% n=760) compared to general health policies (33.51%, n=383, giving a guideline-to-policy citation ratio of 1.98:1. This reflects the requirement for clinical practice guidelines to support recommendations with reference to high-quality evidence.

**Table 5.** The division between Clinical Guidelines and Health Policies.

| Document Classification | Total Citations (n) | Proportion of Citations (%) |
| --- | --- | --- |
| Clinical Guidelines | 760 | 66.49% |
| Health Policies | 383 | 33.51% |
| Total Citations | 1,143 | 100.00% |

The geographic origin of health policy and guidelines was tracked using the issuing organisation metadata in BMJ Impact Analytics. While there are 1,143 total citations across clinical guidelines and policies (Table 5), geographic provenance could only be determined for 704 citations (Table 6). The discrepancy reflects documents geographic metadata was absent or unresolved.

**Table 6.** Geographic distribution of issuing organizations and policy document origins (N=704)

| Country /<br>Regional Grouping | Countries Included | Total<br>Citations<br>(n) | % of Total<br>(N=704) |
| --- | --- | --- | --- |
| UK (Domestic) | United Kingdom (327) | 327 | 46.45% |
| High-Income<br>Anglophone<br>(International) | United States (116), Australia (41),<br>Canada (40), Ireland (24), New<br>Zealand (9) | 230 | 32.67% |
| European / Non-Anglophone High-Income | Belgium (19), Spain (16), Germany<br>(14), Italy (10), Norway (8), France (6),<br>Switzerland (6), Netherlands (5),<br>Sweden (4), Portugal (2), Finland (2) | 92 | 13.07% |
| International /<br>Global Bodies | International organizations (e.g.,<br>WHO, guidelines consortia) (34) | 34 | 4.83% |
| Rest of World /<br>Other Jurisdictions | Japan (7), Brazil (6), Saudi Arabia (2),<br>Malaysia (2), South Korea (1), Chile<br>(1), Colombia (1), Turkey (1) | 21 | 2.98% |
| Total |  | 704 | 100.00% |

The geographic spread of health policy and clinical guideline citations is detailed in Table 6. There is a clear uptake in UK (n=327, 46.45%) and anglophone systems (n=230, 32.67%).

Outside the Anglophone systems, European nations represented 13.07% (n=92) of policy citations, led by Belgium, Spain, Germany, and Italy, while international policy bodies, such as the World Health Organization (WHO) and multinational guideline consortia contribute 4.83% (n=34).

The lowest uptake is from the rest of the world which represent both developed and developing health systems and countries with a strong research culture in prehospital care (Japan and South Korea).

## Discussion

While cross-disciplinary or whole-database analyses report baseline policy citation rates of under 5% to 10%, targeted evaluations in applied health disciplines routinely identify substantially higher policy citation rates. The 23.48% policy citation rate observed in this study is at the lower end of findings reported in similar studies.

Given that this sample originates from clinical practitioners and researchers working in UK NHS ambulance services, rather than researchers based wholly in academia, a higher degree of policy uptake might be anticipated. While translational impact into clinical practice guidelines and health policy is taking place, this finding indicates the need to look at mechanisms that facilitate this and actions to accelerate this translation.

From the top 8 papers in this study, we discover that trials with high citations accrue high policy citation rates. While quality improvement, service evaluations and design and workforce studies have lower citation rates in comparison but high policy citation rates. This potentially points in two directions. Policy citation rates measure something that is different and valuable to certain forms of research and the potential translational value of some studies to clinical guidelines and health policies.

Analysis of the eight most highly cited publications reveals two distinct translational profiles. Large-scale randomised controlled trials and clinical trials accrued both high citation counts and high policy citation rates, reflecting their dual role in advancing clinical research and generating high-quality evidence for clinical guidelines.

Studies addressing service design, workforce models, triage pathways, and quality improvement attracted substantially fewer citations while achieving strong policy citation rates. This highlights two important considerations. First, policy citation rates capture an applied measure of impact that traditional citations miss. Second, it shows that applied, operational and service-level paramedic and ambulance research carries high translational value for decision-makers and clinical guidelines, even where it generates a smaller scholarly impact.

The 1.98:1 ratio of clinical guidelines to health policy citations reflects the need to base clinical decision-making on high-quality evidence. Additionally, clinical guideline panels are typically composed of clinicians and researchers with a grounding in systematic review methodologies and scholarly publication. Decision makers who commission or write health policy operate in a broader context, drawing on diverse informational sources including service evaluations, organizational case studies, public consultations and grey literature. These operational sources frequently lack DOI indexing or fall outside the normal systems of scholarly publication. The sources for policy documents are challenging to measure and represent an area for future research.

The distribution of citations beyond the UK demonstrates clear international reach, with 53.55% (n=377) of all citations originating outside the UK compared with 46.45% (n=327) domestically. However, international citations are heavily concentrated, with Anglophone countries accounting for 32.67% (n=230) and bringing the combined Anglophone share to 79.12%. This concentration likely reflects both a shared language and Anglo-American prehospital care models, where paramedic-led services make UK service designs and triage protocols directly applicable.

The 13.07% (n=92) uptake by non-Anglophone European countries indicates that clinical trial evidence crosses borders regardless of the delivery model deployed. While international bodies account for a relatively small volume, their guidance acts as a translational multiplier, cascading evidence into regional protocols globally. Finally, these geographic patterns should be interpreted alongside the established English-language indexing bias inherent to international policy-tracking databases such as BMJ Impact Analytics.

Translating prehospital research into health policy and clinical guidelines depends on discoverability and access. Institutional repositories such as *amber* provide essential routes to green open access, preserving and disseminating NHS ambulance research across the sector including reports and service evaluations that might otherwise remain inaccessible grey literature.

Beyond digital access, researchers embedded within ambulance services are uniquely placed to showcase their findings to clinical teams, trust boards and national leadership bodies such as the Association of Ambulance Chief Executives (AACE).

To maximise uptake further, authors should ensure that concrete recommendations for practice are explicitly identified in their published research text so they can be readily integrated into policy documents.

More broadly, research confirms that older research outputs tend to accrue more citations over time as evidence consolidates (Szomszor & Adie, 2022). However, clinical guidelines and health policy benefit most from recent research that reflects current clinical realities and operational models. Relying on passive diffusion while waiting for findings to be discovered by clinicians and policy makers is sub-optimal; this creates a clear responsibility for authors to actively promote their work. Healthcare libraries also play an essential role in bridging this gap by identifying, synthesizing and foregrounding current prehospital evidence through their targeted evidence and current awareness services.

### Recommendations

- Active evidence promotion: Authors and ambulance services researchers should proactively share findings across through their organisational networks and not rely on passive academic discovery.
- Explicit clinical takeaways: Authors should write structured practice
- recommendations in published articles to allow guideline development panels to readily identify translational relevance.
- Green open access uptake: Researchers should consistently deposit post-prints and accepted manuscripts into subject repositories such as *amber* to remove paywall barriers for guideline panels and frontline clinicians.
- Healthcare library integration: Ambulance services should embed healthcare library teams into research workflows to support evidence synthesis, horizon scanning and active dissemination of emerging prehospital literature.
- Persistent identifier assignment: Ambulance services should assign DOIs or equivalent persistent identifiers to substantial service evaluations, research reports and grey literature outputs to enable automated tracking across policy databases. In addition, consider using a systematic citation system such as Harvard to further facilitate identification.

### Strengths and limitations

This research paper adheres to the BIBLIO Guidelines (Montazeri et al., 2023) for reporting bibliometric research. The research has several limitations. It relies entirely on locating citations in BMJ Impact Analytics using Digital Object Identifiers. This approach largely ignores reports and grey literature which do not have persistent identifiers.

This research paper adheres to the BIBLIO Guidelines (Montazeri et al., 2023) for reporting bibliometric research. By evaluating the holdings from the *amber* repository, this analysis captures research produced directly by NHS ambulance service personnel rather than relying on broader medical subject groupings.

The research has several limitations. It relies entirely on locating citations in BMJ Impact Analytics using Digital Object Identifiers (DOIs). This approach under represents reports, internal trust audits and grey literature that lack persistent identifiers. Additionally, geographic tracking was limited by documents where issuing organisation metadata could not be resolved.

## Ethics Statement

This research did not require ethics approval.

## Data & Code Availability

The datasets generated and analysed during the current study are available from the corresponding author upon reasonable request.

## Author Contributions

This paper is based on research for a poster presented at the Royal College of Paramedic Research Conference (Holland et al. 2026). Matt Holland is responsible for the methodology, analysis, writing and editing of this paper.

## Use of Artificial Intelligence

During the preparation of this manuscript, the author used large language model assistance (Google Gemini) to assist with copyediting, language refinement and formatting of draft sections. The author reviewed, edited and verified all content and takes full responsibility for the integrity and factual accuracy of the manuscript.

