## Supplemental Table 1 for "Measuring the policy citation rate of United Kingdom (UK) paramedic and ambulance service research: a bibliometric evaluation using BMJ Impact Analytics and the *amber* repository"

**Supplementary File 1: BiBLio Reporting Checklist**

**Title:** Measuring the policy citation rate of United Kingdom (UK) paramedic and ambulance service research: a bibliometric evaluation using BMJ Impact Analytics and the amber repository

**Author:** Matt Holland

**Reporting Guideline:** Preliminary guideline for reporting bibliometric reviews of the biomedical literature (BIBLIO) (Montazeri et al., 2023)

| Section | Item # | BiBLio Checklist Item | Reported? | Location in Manuscript |
|---|---|---|---|---|
| **Title** | 1 | **Identification:** Identify the report as a bibliometric review/analysis in the title | Yes | Title page |
| | 2 | **Issues/topics:** Indicate key topics under investigation and time period coverage | Yes | Title page & **Background** (2006–2026) see page 2 and **Methods** page 4. |
| **Abstract** | 3 | **Structured summary:** Provide a structured abstract (Context, Methods, Findings, Conclusion) | Yes | **Abstract** see page 1. |
| **Introduction** | 4 | **Justification / Rationale:** Present review of existing knowledge and evidence gap | Yes | **Background**, see page 2 and (**Tables 1 & 2**) pages 2 and 3. |
| | 5 | **Objectives:** State explicit research aims or questions | Yes | **Background**, bulleted statement of aims see page 4. |
| **Methods** | 6 | **Search engines (Data sources):** Describe all information sources | Yes | **Methods,** *amber* & BMJ Impact Analytics, see page 4. |

| | | | | |
|---|---|---|---|---|
| | | and platforms searched | | |
| | 7 | **Search strategy:** Provide full search strategy/queries, dates, and retrieval workflow | Yes | **Methods** (June 2026 extraction) see page 4. |
| | 8 | **Data refinement / selection:** Describe screening, filtering, and deduplication process | Yes | **Methods,** DOI extraction & deduplication), see page 4. |
| | 9 | **Eligibility criteria:** Describe inclusion/exclusion criteria, dates, and document types | Yes | **Methods,** NHS ambulance affiliation, 2006–2026, DOIs required, see page 4. |
| | 10 | **Data extraction:** Describe the procedure for extracting and compiling data | Yes | **Methods**, Excel extraction of guidelines, policies, geography, see page 4. |
| | 11 | **Quality assessment (Optional):** Mention quality appraisal of sources if conducted | N/A | Not applicable for citation tracking |
| | 12 | **Data synthesis:** Describe methods and software used for summarizing, tabulations, or displays | Yes | Methods, paragraph 3 Microsoft Excel descriptive bibliometrics, see page 4. |
| **Results** | 13 | **Descriptive statistics:** Report details of search yields, total counts, | Yes | **Results,** Table 3 citation tiers, see pages 4-7. |

| | | | | |
|---|---|---|---|---|
| | | and document distributions | | |
| | 14 | **Schematic map and trend:** Present visual displays, top journals, or citation tiers | Yes | **Table 3** tiers & **Table 4** top 8 highly cited publications, see pages 5-6. |
| | 15 | **Tabulation and summarizing:** Present separate tables summarizing findings by sub-topics | Yes | T**ables 3, 4, 5** & **6**, citation rates, top papers, document types, geography, see pages 5-7. |
| | 16 | **Synthesis of findings:** Synthesize findings and explain observed patterns or differences | Yes | **Results** and **Discussion**, see pages 4-9. |
| **Discussion** | 17 | **Explanations for outcomes:** Provide explanations for observed outcomes, similarities, and differences | Yes | **Discussion**, benchmarks, RCTs vs service design, 1.98:1 ratio, see pages 7-9. |
| | 18 | **Implications / Recommendations:** Discuss implications for policy, practice, or future research | Yes | **Discussion** and **Recommendations**, see pages 7-10. |
| | 19 | **Strengths and limitations:** Discuss strengths and limitations of the review | Yes | **Discussion**, **Strengths** and **Limitations**, see pages 7-10. |
| | 20 | **Conclusion(s):** Provide a general interpretation of results with respect to objectives | Yes | **Abstract**, **Discussion** and **Conclusion** see pages 1, 7-9. |
